# Efficacy and safety of endovascular treatment for acute ischemic stroke due to medium and distal vessel occlusion: a protocol for a systematic review and meta-analysis

**DOI:** 10.1101/2025.09.07.25334910

**Authors:** Eric Dunne, Mehala Subramaniapillai, Areeb Jafrani, Darsh Shah, Andrew Tang, William Tang, Aaron Wen, Snigdha Widge, Aravind Ganesh, Ashkan Shoamanesh, Aristeidis Katsanos, Luciana Catanese, Rachel Couban, Gordon Guyatt, Jason Busse, Arnav Agarwal

## Abstract

**Introduction:** Endovascular thrombectomy (EVT) improves outcomes in large-vessel occlusion (LVO) stroke and has recently shown benefit in dominant M2 occlusions. However, its role in medium vessel occlusions (MeVO) and distal vessel occlusions remains uncertain. This systematic review and meta-analysis evaluates the efficacy and safety of EVT plus usual care versus usual care alone in acute ischemic stroke due to MeVO or distal vessel occlusion.

**Methods:** We will search four electronic databases (Ovid MEDLINE, Embase, CINAHL and Cochrane CENTRAL) from inception to May 2025 without language and other restrictions. Eligible studies will be randomized trials comparing EVT plus medical therapy versus medical therapy alone in adults with acute ischemic stroke due to MeVO or distal vessel occlusion. Paired reviewers will independently screen identified hits for eligibility, extract data from eligible studies, and assess risk of bias using the Risk Of Bias instrument for Use in SysTematic reviews-for Randomised Controlled Trials (ROBUST-RCT). Clinically important outcomes will include functional disability, mortality, neurological function and cognition, quality of life, recanalization, and adverse events. Certainty of evidence will be assessed using Grading of Recommendations, Assessment, Development and Evaluation (GRADE). Random-effects meta-analyses will be conducted, with pre-defined subgroup analyses planned.

**Ethics and Dissemination:** No ethics approval is required. Results will be disseminated via peer-reviewed publication and conference presentations to inform clinicians, guideline developers, and health system decision-makers.

## INTRODUCTION

Endovascular thrombectomy (EVT), when added to medical management, improves functional outcomes among people with acute ischemic stroke caused by large-vessel occlusion (LVO) of the internal carotid artery,^1^ M1 segment of the middle cerebral artery (MCA) or basilar artery.^2–4^ More recently, EVT has also shown benefit in people with occlusion of the dominant M2 segment of the MCA, further expanding its indications.^5^

The role for EVT in adults with occlusions of medium and distal cerebral vessels – including non-dominant M2, M3 and M4 branches of the MCA, branches of the anterior cerebral artery (ACA) and posterior cerebral artery (PCA), and other distal cortical branches – is less well defined. While such strokes tend to involve smaller ischemic volumes and have a generally more favorable natural history compared to LVO strokes, approximately half of these people do not achieve excellent outcomes despite optimal medical management, and a significant proportion remain functionally dependent at 90 days.^6^ Intravenous thrombolysis with alteplase or tenecteplase offers partial benefit, but early recanalization rates remain suboptimal, occurring in fewer than 50% of cases.^7,8^

Post-hoc analyses of large EVT trials and observational data have suggested that EVT may improve outcomes for people with acute ischemic stroke due to medium-vessel occlusions (MeVO) or distal-vessel occlusions.^9,10^ However, until recently, prospective randomized controlled trials (RCTs) directly assessing the efficacy and safety of EVT in this patient population were limited.

Existing international practice guidelines, including those from the American Heart Association/American Stroke Association (AHA/ASA) and the European Stroke Organization (ESO), provide strong recommendations for EVT in LVO, but neither organization definitively recommends for or against EVT for MeVO or distal vessel occlusions, citing insufficient evidence.^11,12^ Insufficient RCT evidence and a lack of definitive practice guidance has led to substantial practice variation internationally. Surveys of stroke centers have highlighted notable heterogeneity in patient selection criteria for EVT in MeVO, particularly regarding therapeutic time windows and considerations of baseline patient functional status, underscoring the urgent need for standardized approaches as ongoing trials aim to address these evidence gaps.^13,14^

In February 2025, the publication of two landmark RCTs – the Endovascular Treatment to Improve Outcomes for Medium Vessel Occlusions (ESCAPE-MeVO) trial and EnDovascular therapy plus usual care versus usual care alone for medIum distal veSsel occlusion sTroke (DISTAL) trial – and press release of a third landmark RCT (DISCOUNT) served as landmark evidence addressing this knowledge gap.^15,16^

Prior low certainty evidence and lack of clear international practice guidance, combined with newly available practice-changing evidence, underscore the need for an updated systematic review and meta-analysis. This systematic review and meta-analysis summarizes the efficacy and safety of EVT added to usual care, compared to usual care alone, in adults with acute ischemic stroke due to MeVO or distal-vessel occlusion.

## METHODS AND ANALYSIS

The protocol for this systematic review adheres to Preferred Reporting Items for Systematic Reviews and Meta-Analyses (PRISMA)-Protocols standards. We will adhere to the PRISMA checklist for reporting our systematic review and meta-analysis. Our protocol will also be registered on PROSPERO.

### Search strategy

With the help of a medical librarian, we will conduct a systematic search of the published literature in four electronic databases from inception to present: Ovid MEDLINE (1946–May 2025), Embase (1947– May 2025), CINAHL (1982– May 2025) and the Cochrane Central Register of Controlled Trials (1991– May 2025). We may update our search closer to the time of publication to capture major landmark trials should they be published in the interim or for which data is otherwise made available.

Search terms will include a combination of keywords and Medical Subject Heading (MeSH) terms: acute ischemic stroke, cerebral infarction, medium vessel occlusion, distal vessel occlusion, M2, M3, M4, anterior cerebral artery, posterior cerebral artery, cortical branch occlusion, non-large vessel occlusion, endovascular therapy, endovascular thrombectomy, mechanical thrombectomy, catheter-based thrombectomy, retrievable stent, embolectomy, clot retrieval, randomized controlled trial. The full search strategy is included as an Appendix I.

We will not apply language or other restrictions in our literature search.

We will supplement our search by reviewing bibliographies of review articles and eligible trials to identify additional studies.

### Eligibility criteria

Eligible studies will be parallel-arm RCTs including adults aged 18 years of age or older with acute ischemic stroke caused by imaging-confirmed occlusion of medium or distal cerebral vessels presenting within 24 hours since they were last seen well and with disabling deficits. For inclusion, studies will report on at least one pre-specified clinically important outcome.

The primary efficacy outcome is functional disability, measured using the modified Rankin scale [mRS] score, reported at 90 days or at longest reported follow-up. Scores range from 0 to 6, with higher scores indicating more severe disability.

Secondary efficacy outcomes of interest are:

1. Hospital length of stay (measured at 90 days or longest reported follow-up).
2. Need for admission to intensive care unit (measured at 90 days or longest reported follow-up).
3. Severity of neurological deficit (measured using the National Institutes of Health Stroke Scale or an alternate scoring system, reported within 48 +/- 24 hours).
4. Cognitive function (measured using the Montreal Cognitive Assessment or an alternative scoring system, reported at 90 days or longest reported follow-up)
5. Health-related quality of life (measured using the EuroQol 5-Dimension 5-Level Index or an alternative scoring system, reported at 90 days or longest reported follow-up)

The primary safety outcome is symptomatic intracranial hemorrhage, reported at 48 +/- 24 hours.

Secondary safety outcomes of interest are:

1. All-cause death (measured at 90 days or longest reported follow-up).
2. Serious adverse events (reported at 90 days or longest reported follow-up).
3. Radiological intracranial hemorrhage (reported at 48 +/- 24 hours), based on Heidelberg intracranial hemorrhage classification.
4. Stroke progression (reported at 90 days or longest reported follow-up)
5. Iatrogenic arterial perforation (reported at 90 days or longest reported follow-up).
6. Iatrogenic vessel dissection (reported at 90 days or longest reported follow-up).
7. Major extracranial bleeding (including access site or retroperitoneal hematoma) (reported at 90 days or longest reported follow-up).
8. Access site neuropathy (reported at 90 days or longest reported follow-up).
9. Embolization to previously unaffected territory (reported at 90 days or longest reported follow-up).
10. Other specific clinically important harms (reported in at least two or more trials) (reported at 90 days or longest reported follow-up).

If available, we will also collect and report data related to recanalization and/or reperfusion of occluded vessels.

We will use trial definitions for all outcomes.

We will include articles meeting eligibility criteria, irrespective of language and publication status; specifically, we will include grey literature and conference abstracts.

### Study selection

Using pre-tested standardized screening forms with accompanying instructions and following completion of a pilot exercise, paired reviewers will screen identified hits at title-and-abstract and full-text levels. Reviewers will resolve disagreements by discussion, with adjudication by a third reviewer when necessary. We will use Covidence (https://covidence.org/) for screening and study selection. Reasons for exclusion at the full-text screening level will be summarized using a PRISMA flow diagram.

### Data extraction and risk of bias assessment

Paired reviewers will independently extract data and assess risk of bias for each included study at the outcome level using pre-tested, standardized extraction forms with accompanying guidance. Discrepancies will be resolved through discussion and, if necessary, adjudication by a third reviewer.

We will extract data related to the study population, interventions, comparators, and outcomes of interest. For the population, the following variables will be collected: geographic region; care setting (e.g., intensive care unit vs. non-intensive care unit); baseline stroke severity (measured based on NIHSS); Alberta Stroke Program Early CT Score (ASPECTS); sex; age; race; and relevant medical comorbidities, including hypertension, dyslipidemia, ischemic heart disease, diabetes mellitus, prior stroke or transient ischemic attack, smoking status, atrial fibrillation, heart failure, peripheral arterial disease, and other cardiovascular conditions or risk factors. Additional variables will include time intervals from last known well to randomization, imaging, and arterial puncture; use of intravenous thrombolysis; occlusion location; baseline imaging findings (e.g., early ischemic changes, tandem occlusion); and final Medium Vessel Occlusion-expanded Thrombolysis in Cerebral Ischemia (MeVO-eTICI) reperfusion scores.

Risk of bias will be evaluated using the ROBUST-RCT tool^17^, which assesses bias across the following domains: random sequence generation (selection bias), allocation concealment (selection bias), blinding of participants and personnel (performance bias), blinding of outcome assessment (detection bias), incomplete outcome data (attrition bias), selective outcome reporting (reporting bias), and other potential sources of bias (e.g., early trial termination, influence of funding). Each domain is evaluated using a series of signaling questions, which guide reviewers in forming a risk judgment of low risk, some concerns, or high risk of bias.

### Assessment of certainty of evidence

We will assess the overall certainty of evidence for each pre-specified clinically important outcome using the Grading of Recommendations Assessment, Development and Evaluation (GRADE) approach. This assessment will be based on five domains: risk of bias, imprecision, inconsistency, indirectness, and publication bias. Overall certainty of evidence will be rated as high, moderate, low, or very low. We will consider downgrading for risk of bias in the context of lack of blinding for subjective outcomes only. Issues related to missing data will be addressed within the risk of bias assessments as per ROBUST-RCT. For judgements regarding imprecision, we will examine whether the confidence intervals around effect estimates cross either the null or pre-defined minimally important difference (MID) thresholds specific to clinically important outcomes.

### Statistical analysis

We will perform random-effects meta-analyses using the inverse variance method. For dichotomous outcomes, pooled relative effects will be expressed as risk ratios (RRs) with corresponding 95% confidence intervals (CIs). Absolute effect estimates will be derived using pooled event rates from the control arms of included studies to inform baseline risks. For continuous outcomes, pooled effects will be reported as mean differences (MDs) with corresponding 95% CIs. We will assume a normal distribution for continuous data and will convert interquartile ranges (IQRs) to standard deviations (SDs) using methods recommended by the Cochrane Collaboration. We will assess heterogeneity across studies using the χ² test for homogeneity, the I² statistic, and visual inspection of forest plots.

If sufficient data are available, we will plan the following subgroup analyses:

- Time from last seen well to treatment (hypothesis: increased effect of EVT with decreased time from last seen well).
- Stroke severity (i.e. baseline NIHSS) (hypothesis: increased efficacy of EVT among those with higher baseline NIHSS)
- Occlusion site (M2, M3 or M4, anterior cerebral artery, posterior cerebral artery) (hypothesis: increased effect of EVT for M2 and PCA compared to M3 or M4 and ACA)
- Pre-procedure IV thrombolysis (hypothesis: increased effect of EVT without concomitant thrombolysis)

We will rate the credibility of effect modification analyses using the Instrument for assessing the Credibility of Effect Modification Analyses (ICEMAN) tool.^18^

We will perform a sensitivity analysis limiting to studies at low risk of bias.

## ETHICS AND DISSEMINATION

As this study is a systematic review of previously published data, ethics approval is not required. We intend to disseminate our findings through presentations at national and international medical conferences. In addition, we plan to submit the final manuscript for publication in a peer-reviewed journal that is widely read by general practitioners, internists, and other healthcare professionals involved in stroke care.

## PATIENT AND PUBLIC INVOLVEMENT

The research question and study design were developed with a focus on patient-important outcomes. Patients and members of the public were not directly involved in the design of this review.

## Data Availability

All data produced in the present study are available upon reasonable request to the authors.

## Appendix I - Summary of search and strategies Endovascular Treatment for occlusive stroke

May 6, 2025

|  |  |
| --- | --- |
| MEDLINE | 1547 |
| Embase | 2937 |
| CENTRAL | 636 |
| CINAHL | 285 |
| Subtotal | 5405 |
| -duplicates | -1317 |
| Total | 4088 |

### Ovid MEDLINE(R) ALL <1946 to May 05, 2025>

1. exp Stroke/191791
2. exp brain infarction/ or exp cerebral infarction/ 45119
3. (stroke adj3 (acute or ischemic or ischaemi* or mild)).mp. 113728
4. (cerebr* adj3 (infarct* or accident)).mp. 61755
5. (brain* adj3 (ischemi* or ischaemi*)).mp. [mp=title, book title, abstract, original title, name of substance word, subject heading word, floating sub-heading word, keyword heading word, organism supplementary concept word, protocol supplementary concept word, rare disease supplementary concept word, unique identifier, synonyms, population supplementary concept word, anatomy supplementary concept word] 86492
6. or/1-5 290792
7. thrombectomy/ or mechanical thrombolysis/ 13281
8. Endovascular Procedures/ 32019
9. (thrombectomy or thrombolysis or endovascular or intravascular).mp. [mp=title, book title, abstract, original title, name of substance word, subject heading word, floating sub-heading word, keyword heading word, organism supplementary concept word, protocol supplementary concept word, rare disease supplementary concept word, unique identifier, synonyms, population supplementary concept word, anatomy supplementary concept word] 185547
10. (embolectomy* or “stent retriever” or “stent-retriever” or “aspiration”).mp. [mp=title, book title, abstract, original title, name of substance word, subject heading word, floating sub-heading word, keyword heading word, organism supplementary concept word, protocol supplementary concept word, rare disease supplementary concept word, unique identifier, synonyms, population supplementary concept word, anatomy supplementary concept word] 109723
11. (Stentriever or ADAPT or “A direct aspiration, first pass technique” or Direct thrombus aspiration or Direct aspiration or Stents or catheter or microcatheter or neuroangiography or Medtronic Solitaire or Medtronic Mindframe Capture or Balt Catch or Gateway or Penumbra or Trevo ProVue or Trevo XP or Covidien Solitaire or Revive SE or Concentric Merci or Penumbra system).mp. [mp=title, book title, abstract, original title, name of substance word, subject heading word, floating sub-heading word, keyword heading word, organism supplementary concept word, protocol supplementary concept word, rare disease supplementary concept word, unique identifier, synonyms, population supplementary concept word, anatomy supplementary concept word] 389794
12. clot retrieval.mp. 260
13. or/7-11 633805
14. 6 and 13 37910
15. (occlusion* or occlude*).mp. [mp=title, book title, abstract, original title, name of substance word, subject heading word, floating sub-heading word, keyword heading word, organism supplementary concept word, protocol supplementary concept word, rare disease supplementary concept word, unique identifier, synonyms, population supplementary concept word, anatomy supplementary concept word] 259195
16. 14 and 15 12733
17. exp Cerebral Arteries/ 30552
18. (“distal vessel” or “medium vessel” or “DMVO” or “MEVO” or “ACA” or “anterior cerebral artery” or “PCA” or “posterior cerebral artery” or “MCA” or “middle cerebral artery” or “PICA” or “posterior inferior cerebellar artery” or “AICA” or “anterior inferior cerebellar artery” or “SCA” or “superior cerebellar artery” or “posterior circulation” or M2 or M3 or M4 or M5 or P1 or P2 or P3 or A1 or A2 or A3 or mild).mp. 1093681
19. 17 or 18 1109885
20. 16 and 19 5489
21. randomized controlled trial.pt. 639582
22. controlled clinical trial.pt. 95697
23. randomi?ed.ab. 826901
24. placebo.ab. 258908
25. drug therapy.fs. 2820299
26. randomly.ab. 460852
27. trial.ab. 755656
28. groups.ab. 2858653
29. or/21-28 6327867
30. exp animals/ not humans.sh. 5346533
31. 29 not 30 5548361
32. 20 and 31 1547

### Embase (OVID)

Database: Embase <1974 to 2025 May 05> Search Strategy:

1. exp cerebrovascular accident/ (489821)
2. exp brain infarction/ (98729)
3. (stroke adj3 (acute or ischemic or ischaemi* or mild)).mp. (196005)
4. (cerebr* adj3 (infarct* or accident)).mp. (489735)
5. or/1-4 (631383)
6. exp thrombectomy/ or mechanical thrombectomy/ (47341)
7. exp endovascular surgery/ (58838)
8. (thrombectomy or thrombolysis or endovascular or intravascular).mp. (304906)
9. (embolectomy* or “stent retriever” or “stent-retriever” or “aspiration”).mp. (252076)
10. (Stentriever or ADAPT or “A direct aspiration, first pass technique” or Direct thrombus aspiration or Direct aspiration or Stents or catheter or microcatheter or neuroangiography or Medtronic Solitaire or Medtronic Mindframe Capture or Balt Catch or Gateway or Penumbra or Trevo ProVue or Trevo XP or Covidien Solitaire or Revive SE or Concentric Merci or Penumbra system).mp. (611024)
11. clot retrieval.mp. (604)
12. or/6-11 (1080834)
13. 5 and 12 (87115)
14. (occlusion* or occlud* or occlusive).mp. [mp=title, abstract, heading word, drug trade name, original title, device manufacturer, drug manufacturer, device trade name, keyword heading word, floating subheading word, candidate term word] (522334)
15. 13 and 14 (30283)
16. exp brain artery/ (72384)
17. (“distal vessel” or “medium vessel” or “DMVO” or “MEVO” or “ACA” or “anterior cerebral artery” or “PCA” or “posterior cerebral artery” or “MCA” or “middle cerebral artery” or “PICA” or “posterior inferior cerebellar artery” or “AICA” or “anterior inferior cerebellar artery” or “SCA” or “superior cerebellar artery” or “posterior circulation” or M2 or M3 or M4 or M5 or P1 or P2 or P3 or A1 or A2 or A3 or mild).mp. (1522157)
18. 16 or 17 (1545472)
19. 15 and 18 (12887)
20. randomized controlled trial/ (879504)
21. Controlled clinical study/ (445462)
22. random$.ti,ab. (2201674)
23. randomization/ (100708)
24. intermethod comparison/ (313952)
25. placebo.ti,ab. (392267)
26. (compare or compared or comparison).ti. (650041)
27. ((evaluated or evaluate or evaluating or assessed or assess) and (compare or compared or comparing or comparison)).ab. (3130359)
28. (open adj label).ti,ab. (123542)
29. ((double or single or doubly or singly) adj (blind or blinded or blindly)).ti,ab. (293913)
30. double blind procedure/ (231548)
31. parallel group$1.ti,ab. (35383)
32. (crossover or cross over).ti,ab. (133724)
33. ((assign$ or match or matched or allocation) adj5 (alternate or group$1 or intervention$1 or patient$1 or subject$1 or participant$1)).ti,ab. (458263)
34. (assigned or allocated).ti,ab. (542074)
35. (controlled adj7 (study or design or trial)).ti,ab. (502657)
36. (volunteer or volunteers).ti,ab. (297832)
37. human experiment/ (690155)
38. trial.ti. (456137)
39. or/20-38 (6969166)
40. (random$ adj sampl$ adj7 (“cross section$” or questionnaire$1 or survey$ or database$1)).ti,ab. not (comparative study/ or controlled study/ or randomi?ed controlled.ti,ab. or randomly assigned.ti,ab.) (10423)
41. Cross-sectional study/ not (randomized controlled trial/ or controlled clinical study/ or controlled study/ or randomi?ed controlled.ti,ab. or control group$1.ti,ab.) (440205)
42. (((case adj control$) and random$) not randomi?ed controlled).ti,ab. (23515)
43. (Systematic review not (trial or study)).ti. (323258)
44. (nonrandom$ not random$).ti,ab. (20153)
45. “Random field$”.ti,ab. (3151)
46. (random cluster adj3 sampl$).ti,ab. (1746)
47. (review.ab. and review.pt.) not trial.ti. (1282328)
48. “we searched”.ab. and (review.ti. or review.pt.) (57806)
49. “update review”.ab. (153)
50. (databases adj4 searched).ab. (75924)
51. (rat or rats or mouse or mice or swine or porcine or murine or sheep or lambs or pigs or piglets or rabbit or rabbits or cat or cats or dog or dogs or cattle or bovine or monkey or monkeys or trout or marmoset$1).ti. and animal experiment/ (1299018)
52. Animal experiment/ not (human experiment/ or human/) (2738626)
53. or/40-52 (4808838)
54. 39 not 53 (6120109)
55. 19 and 54 (2937)

### Cochrane Library (Wiley)

Search Name:

Date Run: 07/05/2025 15:44:25 Comment: ID Search Hits

#1 MeSH descriptor: [Stroke] explode all trees 17959

#2 MeSH descriptor: [Brain Infarction] explode all trees 1942

#3 (stroke NEAR/3 (acute or ischemic or ischaemi* or mild)) 22214 #4 (cerebr* NEAR/3 (infarct* or accident)) 24566

#5 (brain* NEAR/3 (ischemi* or ischaemi*)) 7594 #6 #1 or #2 or #3 or #4 or #5 49724

#7 MeSH descriptor: [Thrombectomy] explode all trees 836

#8 MeSH descriptor: [Mechanical Thrombolysis] explode all trees 73

#9 thrombectomy or thrombolysis or endovascular or intravascular 17929

#10 embolectomy* or “stent retriever” or “stent-retriever” or “aspiration” 12826

#11 Stentriever or ADAPT or “A direct aspiration, first pass technique” or Direct thrombus aspiration or Direct aspiration or Stents or catheter or microcatheter or neuroangiography or Medtronic Solitaire or Medtronic Mindframe Capture or Balt Catch or Gateway or Penumbra or Trevo ProVue or Trevo XP or Covidien Solitaire or Revive SE or Concentric Merci or Penumbra system. 46049

#12 clot retrieval 147

#13 #7 or #8 or #9 or #10 or #11 or #12 70743 #14 occlusion* or occlude* or occlusive 24119 #15 #6 and #13 and #14 2263

#16 MeSH descriptor: [Cerebral Arteries] explode all trees 665

#17 “distal vessel” or “medium vessel” or “DMVO” or “MEVO” or “ACA” or “anterior cerebral artery” or “PCA” or “posterior cerebral artery” or “MCA” or “middle cerebral artery” or “PICA” or “posterior inferior cerebellar artery” or “AICA” or “anterior inferior cerebellar artery” or “SCA” or “superior cerebellar artery” or “posterior circulation” or M2 or M3 or M4 or M5 or P1 or P2 or P3 or A1 or A2 or A3 or mild 174692

#18 #16 or #17 174842

#19 #15 and #18 in Trials 636

### CINAHL (EBSCO)

| Wed, May 7, 2025 3:14:58 PM |  |  |  |  |
| --- | --- | --- | --- | --- |
| # | Query | Limiters/Expanders | Last Run Via | Results |
| S43 | S38 AND S39 AND S42 | Expanders - Apply equivalent subjects<br>Search modes - Proximity | Interface - EBSCOhost<br>Research Databases<br>Search Screen - Advanced Search<br>Database - CINAHL | 285 |
| S42 | S40 OR S41 | Expanders - Apply equivalent subjects<br>Search modes - Proximity | Interface - EBSCOhost<br>Research Databases<br>Search Screen - Advanced Search<br>Database - CINAHL | 139,514 |
| S41 | TX ("distal vessel" or "medium vessel" or "DMVO" or "MEVO" or "ACA" or "anterior cerebral artery" or "PCA" or "posterior cerebral artery" or "MCA" or "middle cerebral artery" or "PICA" or "posterior inferior cerebellar artery" or "AICA" or "anterior inferior cerebellar artery" or "SCA" or "superior cerebellar artery" or "posterior circulation" or M2 or M3 or M4 or M5 or P1 or P2 or P3 or A1 or A2 or A3 or mild)) | Expanders - Apply equivalent subjects<br>Search modes - Proximity | Interface - EBSCOhost<br>Research Databases<br>Search Screen - Advanced Search<br>Database - CINAHL | 136,972 |
| S40 | (MH "Cerebral Arteries+") | Expanders - Apply equivalent subjects<br>Search modes - Proximity | Interface - EBSCOhost<br>Research Databases<br>Search Screen - Advanced Search<br>Database - CINAHL | 5,746 |
| S39 | TX occlusion* OR TX occlude* OR TX occlusive | Expanders - Apply equivalent subjects<br>Search modes - Proximity | Interface - EBSCOhost<br>Research Databases<br>Search Screen - Advanced Search<br>Database - CINAHL | 42,931 |
| S38 | S30 AND S37 | Expanders - Apply equivalent subjects<br>Search modes - Proximity | Interface - EBSCOhost<br>Research Databases<br>Search Screen - Advanced Search<br>Database - CINAHL | 3,332 |
| S37 | S31 OR S32 OR S33 OR S34 OR S35 OR S36 | Expanders - Apply equivalent subjects<br>Search modes - Proximity | Interface - EBSCOhost<br>Research Databases<br>Search Screen - Advanced Search<br>Database - CINAHL | 178,480 |
| S36 | TX clot retrieval | Expanders - Apply equivalent subjects<br>Search modes - Proximity | Interface - EBSCOhost<br>Research Databases<br>Search Screen - Advanced Search<br>Database - CINAHL | 85 |
| S35 | TX (Stentriever or ADAPT or "A direct aspiration, first pass technique" or Direct thrombus aspiration or Direct aspiration or Stents or catheter or microcatheter or neuroangiography or Medtronic Solitaire or Medtronic Mindframe Capture or Balt Catch or Gateway or Penumbra or Trevo ProVue or Trevo XP or Covidien Solitaire or Revive SE or Concentric Merci or Penumbra system) | Expanders - Apply equivalent subjects<br>Search modes - Proximity | Interface - EBSCOhost<br>Research Databases<br>Search Screen - Advanced Search<br>Database - CINAHL | 126,597 |
| S34 | TX (embolectomy* or "stent retriever" or "stent-retriever" or "aspiration") | Expanders - Apply equivalent subjects<br>Search modes - Proximity | Interface - EBSCOhost<br>Research Databases<br>Search Screen - Advanced Search<br>Database - CINAHL | 21,282 |
| S33 | TX (thrombectomy or thrombolysis or endovascular or intravascular) | Expanders - Apply equivalent subjects<br>Search modes - Proximity | Interface - EBSCOhost<br>Research Databases<br>Search Screen - Advanced Search<br>Database - CINAHL | 42,436 |
| S32 | (MH "Endovascular Procedures+") | Expanders - Apply equivalent subjects<br>Search modes - Proximity | Interface - EBSCOhost<br>Research Databases<br>Search Screen - Advanced Search<br>Database - CINAHL | 4,784 |
| S31 | (MH "Thrombectomy") | Expanders - Apply equivalent subjects<br>Search modes - Proximity | Interface - EBSCOhost<br>Research Databases<br>Search Screen - Advanced Search<br>Database - CINAHL | 3,923 |
| S30 | S23 AND S29 | Expanders - Apply equivalent subjects<br>Search modes - Proximity | Interface - EBSCOhost<br>Research Databases<br>Search Screen - Advanced Search<br>Database - CINAHL | 23,776 |
| S29 | S24 OR S25 OR S26 OR S27 OR S28 | Expanders - Apply equivalent subjects<br>Search modes - Proximity | Interface - EBSCOhost<br>Research Databases<br>Search Screen - Advanced Search<br>Database - CINAHL | 105,112 |
| S28 | TX ((stroke N3 (acute or ischemic or ischaemi* or mild))) | Expanders - Apply equivalent subjects<br>Search modes - Proximity | Interface - EBSCOhost<br>Research Databases<br>Search Screen - Advanced Search<br>Database - CINAHL | 37,949 |
| S27 | TX ((brain* N3 (ischemi* or ischaemi*))) | Expanders - Apply equivalent subjects<br>Search modes - Proximity | Interface - EBSCOhost<br>Research Databases<br>Search Screen - Advanced Search<br>Database - CINAHL | 5,370 |
| S26 | TX ((cerebr* N3 (infarct* or accident))) | Expanders - Apply equivalent subjects<br>Search modes - Proximity | Interface - EBSCOhost<br>Research Databases<br>Search Screen - Advanced Search<br>Database - CINAHL | 8,481 |
| S25 | (MH "Cerebral Infarction") | Expanders - Apply equivalent subjects<br>Search modes - Proximity | Interface - EBSCOhost<br>Research Databases<br>Search Screen - Advanced Search<br>Database - CINAHL | 524 |
| S24 | (MH "Stroke+") | Expanders - Apply equivalent subjects<br>Search modes - Proximity | Interface - EBSCOhost<br>Research Databases<br>Search Screen - Advanced Search<br>Database - CINAHL | 82,279 |
| S23 | S22 NOT S21 | Expanders - Apply equivalent subjects<br>Search modes - Proximity | Interface - EBSCOhost<br>Research Databases<br>Search Screen - Advanced Search<br>Database - CINAHL | 1,040,807 |
| S22 | S1 OR S2 OR S3 OR S4 OR S5 OR S6 OR S7 OR S8 OR S9 OR S10 OR S11 OR S12 OR S13 OR S14 OR S15 | Expanders - Apply equivalent subjects<br>Search modes - Proximity | Interface - EBSCOhost<br>Research Databases<br>Search Screen - Advanced Search<br>Database - CINAHL | 1,088,975 |
| S21 | S19 NOT S20 | Expanders - Apply equivalent subjects<br>Search modes - Proximity | Interface - EBSCOhost<br>Research Databases<br>Search Screen - Advanced Search<br>Database - CINAHL | 211,457 |
| S20 | MH (HUMAN) | Expanders - Apply equivalent subjects<br>Search modes - Proximity | Interface - EBSCOhost<br>Research Databases<br>Search Screen - Advanced Search<br>Database - CINAHL | 2,891,365 |
| S19 | S16 OR S17 OR S18 | Expanders - Apply equivalent subjects<br>Search modes - Proximity | Interface - EBSCOhost<br>Research Databases<br>Search Screen - Advanced Search<br>Database - CINAHL | 245,893 |
| S18 | TI (ANIMAL MODEL*) | Expanders - Apply equivalent subjects<br>Search modes - Proximity | Interface - EBSCOhost<br>Research Databases<br>Search Screen - Advanced Search<br>Database - CINAHL | 3,678 |
| S17 | MH (ANIMAL STUDIES) | Expanders - Apply equivalent subjects<br>Search modes - Proximity | Interface - EBSCOhost<br>Research Databases<br>Search Screen - Advanced Search<br>Database - CINAHL | 154,311 |
| S16 | MH ANIMALS+ | Expanders - Apply equivalent subjects<br>Search modes - Proximity | Interface - EBSCOhost<br>Research Databases<br>Search Screen - Advanced Search<br>Database - CINAHL | 100,545 |
| S15 | AB (CLUSTER W3 RCT) | Expanders - Apply equivalent subjects<br>Search modes - Proximity | Interface - EBSCOhost<br>Research Databases<br>Search Screen - Advanced Search<br>Database - CINAHL | 530 |
| S14 | MH (CROSSOVER DESIGN) OR MH (COMPARATIVE STUDIES) | Expanders - Apply equivalent subjects<br>Search modes - Proximity | Interface - EBSCOhost<br>Research Databases<br>Search Screen - Advanced Search<br>Database - CINAHL | 511,161 |
| S13 | AB (CONTROL W5 GROUP) | Expanders - Apply equivalent subjects<br>Search modes - Proximity | Interface - EBSCOhost<br>Research Databases<br>Search Screen - Advanced Search<br>Database - CINAHL | 154,755 |
| S12 | PT (randomized controlled trial) | Expanders - Apply equivalent subjects<br>Search modes - Proximity | Interface - EBSCOhost<br>Research Databases<br>Search Screen - Advanced Search<br>Database - CINAHL | 162,477 |
| S11 | MH (placebos) | Expanders - Apply equivalent subjects<br>Search modes - Proximity | Interface - EBSCOhost<br>Research Databases<br>Search Screen - Advanced Search<br>Database - CINAHL | 14,319 |
| S10 | MH (sample size) AND AB (assigned OR allocated OR control) | Expanders - Apply equivalent subjects<br>Search modes - Proximity | Interface - EBSCOhost<br>Research Databases<br>Search Screen - Advanced Search<br>Database - CINAHL | 4,509 |
| S9 | TI (trial) | Expanders - Apply equivalent subjects<br>Search modes - Proximity | Interface - EBSCOhost<br>Research Databases<br>Search Screen - Advanced Search<br>Database - CINAHL | 206,648 |
| S8 | AB (random*) | Expanders - Apply equivalent subjects<br>Search modes - Proximity | Interface - EBSCOhost<br>Research Databases<br>Search Screen - Advanced Search<br>Database - CINAHL | 416,428 |
| S7 | TI (randomised OR randomized) | Expanders - Apply equivalent subjects<br>Search modes - Proximity | Interface - EBSCOhost<br>Research Databases<br>Search Screen - Advanced Search<br>Database - CINAHL | 161,553 |
| S6 | MH cluster sample | Expanders - Apply equivalent subjects<br>Search modes - Proximity | Interface - EBSCOhost<br>Research Databases<br>Search Screen - Advanced Search<br>Database - CINAHL | 6,080 |
| S5 | MH pretest-posttest design | Expanders - Apply equivalent subjects<br>Search modes - Proximity | Interface - EBSCOhost<br>Research Databases<br>Search Screen - Advanced Search<br>Database - CINAHL | 58,763 |
| S4 | MH random assignment | Expanders - Apply equivalent subjects<br>Search modes - Proximity | Interface - EBSCOhost<br>Research Databases<br>Search Screen - Advanced Search<br>Database - CINAHL | 91,602 |
| S3 | MH single-blind studies | Expanders - Apply equivalent subjects<br>Search modes - Proximity | Interface - EBSCOhost<br>Research Databases<br>Search Screen - Advanced Search<br>Database - CINAHL | 16,315 |
| S2 | MH double-blind studies | Expanders - Apply equivalent subjects<br>Search modes - Proximity | Interface - EBSCOhost<br>Research Databases<br>Search Screen - Advanced Search<br>Database - CINAHL | 54,678 |
| S1 | MH randomized controlled trials | Expanders - Apply equivalent subjects<br>Search modes - Proximity | Interface - EBSCOhost<br>Research Databases<br>Search Screen - Advanced Search<br>Database - CINAHL | 150,121 |

